# Bioinformatic Screen with Clinical Validation for the Identification of Novel Stool Based mRNA Biomarkers for the Detection of Colorectal Lesions Including Advanced Precancerous Lesions

**DOI:** 10.1101/2025.07.16.25331674

**Authors:** Houcong Liu, Loren Hansen, Changpu Song, Haijiu Lin, Dan Chen, Zhufang Chen, Hekai Zhou, Xiao Yang, Wenying Pan, Jihui Du

**Affiliations:** Research Center for Clinical and Translational Medicine, Shenzhen Nanshan People’s Hospital and the 6th Affiliated Hospital of Shenzhen University Medical School, Shenzhen, Guangdong 518052, China; El Capitan Biosciences, 7068 Koll Center Pkwy, Suite 402, Pleasanton, CA 94566; Guangdong Jiyin Biotech, D3 Building, TCL international E city, no. 1001, Zhong Shan Yuan Road, Nanshan District. Shenzhen, China

**Keywords:** mRNA biomarkers, stool, colorectal cancer, diagnostics

## Abstract

Messenger RNA (mRNA) stool based biomarkers represent a promising approach for the diagnosis of colorectal cancer (CRC) and advanced precancerous lesions (AA). But it is unclear which mRNA biomarkers have the most clinical utility. This study aims to partially fill this gap by performing an analysis which first ranks genes based on their expression profile in RNA-seq tissue datasets.

The diagnostic performance of the top 20 ranked genes was then tested using 113 clinical samples (CRC N=33, AA N=28, Controls N=53). Fourteen of the genes had significant differential expression in the stool of CRC patients compared to controls (false discovery rate or FDR < 0.05). The Pearson correlation coefficient between tissue and stool expression was 0.57 (p-value = 0.007). The combined performance of the 20 genes in clinical stool samples had an area under the receiver operator curve (AUC) of 0.94 for CRC detection (sensitivity 75.5%, specificity 95%) and an AUC of 0.83 (sensitivity 55.8%, specificity 92.6%) for AA detection. The ability to use existing public transcriptomic datasets to identify promising candidate genes can substantially reduce the cost and effort required to screen for clinically useful mRNA biomarkers.

## Background

Globally, ColoRectal Cancer (CRC) is the third most commonly diagnosed cancer and the second most common cause of death in cancer-related deaths for both men and women^1^. CRC is therefore a serious global health burden. Colorectal cancer almost always follows the adenoma-carcinoma pathway in which a series of stepwise mutations and epigenetic modifications slowly transform normal epithelium to dysplastic epithelium; this transformation commonly takes 10-15 years ^2^. The slow progression from benign polyps to malign growths provides an opportunity to identify and remove advanced precursor lesions (Advanced Adenoma AA) or early stage malignant tumors. Removal of precursor lesions prevents the development of malignant disease, while the identification and treatment of CRC in the early stages leads to a much improved prognosis. Five-year survival for patients with localized disease is ∼90% but only 10% once the disease has metastasized to distal organs ^3^. While removal of adenomas and advanced adenomas has been shown to reduce CRC incidence rates ^4^.

Given the slow development from benign polyps to malign growths CRC is an attractive target for prevention and early detection through population based screening programs. Indeed a number of countries have implemented some form of population based screening that have led to reductions in CRC incidence and mortality ^5^. Colonoscopies are the gold standard approach to screen for precursor lesions and early stage disease with excellent sensitivity and specificity ^6^. Unfortunately compliance with a colonoscopy is frequently not optimal due to the invasive nature of the procedure, unpleasant preparation, and time requirements ^7^. In addition, colonoscopy resources in developing countries are frequently not sufficient to support population level CRC screening ^8^.

Due to these issues a number of alternative approaches have been developed the most prominent of which is non-invasive stool based tests ^5,9^. The most common stool based test worldwide is detection of occult blood in stool ^2,6,10^. Fecal immunochemical tests (FIT) tests are cost effective, have high sensitivity and specificity in detecting the presence of blood associated proteins in stool and do not require dietary restrictions. Unfortunately occult blood detection using FIT while it has good specificity for CRC/AA detection at ∼90-95% suffers from poor sensitivity in AA detection (10-40%) and modest sensitivity in detection of early stage CRC tumors (37-70%) ^5,6,11,12^.

Given the limitations of occult blood detection, alternate strategies that depend on the exfoliation of tumor cells into the colon-rectal lumen have been developed. The most mature approach is detection of characteristic tumor/AA epigenetic marks plus somatic mutations plus detection of occult blood using FIT. Indeed, there is a FDA-approved multitarget DNA methylation/somatic mutation+FIT test available for screening purposes ^9^. While pairing occult blood biomarkers (FIT) with detection of tumor DNA based biomarkers does increase the sensitivity compared with FIT alone for early stage and AA detection (CRC 92.3%, AA 42%) this comes at the cost of lower specificity (87% specificity versus FIT with ∼95% specificity) and greatly increased cost compared with FIT^13^.

Detection of tumor/AA derived mRNA from exfoliated cells instead of DNA has a number of potential advantages. Aberrant tumor derived DNA will normally consist of at most 2 copies per shed tumor cell while aberrantly expressed mRNA may have many hundreds or thousands of copies per shed tumor cell. The additional detectable signal from RNA based biomarkers could increase the sensitivity of detection for small early stage tumors and advanced adenomas while maintaining good specificity. There have been a number of studies demonstrating that measuring CRC associated mRNA in stool can be a reliable method to detect colorectal cancers and advanced adenomas.^14,15^. Indeed on the basis of a large-scale clinical validation trial in the intended use population the FDA recently approved a mRNA based test for CRC/AA population based screening^16^.

These and other studies have established that mRNA based biomarkers can be an effective method for the detection of colorectal neoplasms and advanced precancerous lesions. However, which mRNA biomarkers are best suited for CRC/AA screening has yet to be identified or validated. Most existing studies have tested a handful of published genes that on the basis of tissue expression have been shown to be associated with CRC. While this has been a productive approach it is clear that not all published CRC associated mRNA biomarkers are suitable for a high performing stool based screening test. It is also clear that selection of mRNA biomarkers is critical in determining test performance^17,18^. Indeed, simply because a gene is over or under expressed in CRC tissue does not necessarily mean that gene’s pattern of expression in tissue is best suited for a stool based CRC screening test.

In addition, cells that are exfoliated into stool are in a very different and stressful environment compared to cells in tissue, therefore it is expected the transcriptional profile of cells shed into stool may differ significantly from the tissue obtained transcriptional profile. What impact this may have is currently unclear.

Here we take advantage of the thousands of high quality publicly available RNA-seq tissue transcriptomics datasets to perform a comprehensive bioinformatic analysis to rank genes that based on their pattern of expression in CRC and normal tissue are most suitable for a stool based CRC screening test. After identification, we tested the top ranked genes on clinical samples and showed that while tissue is not a perfect predictor of stool mRNA representation there is enough overlap that tissue transcriptomics can productively be used to identify stool based mRNA biomarkers that have strong clinical utility.

## Materials and Methods

### Ranking Genes

CRC Transcriptomic datasets and paired normal tissue samples were downloaded from The Cancer Genome Atlas (TCGA) database ^19^ additional healthy colon controls were downloaded from the Genotype-Tissue Expression (GTEx) database. The combined TCGA/GTE dataset consisted of 478 colon cancer tissue samples, and 692 normal colon/rectum tissue samples. The data consisted of gene count matrices.

Batch correction on the downloaded data to merge the healthy controls from the TCGA database with the GTEx normal samples was performed using combat-seq ^20^. A differential expression analysis comparing gene expression in CRC tissue vs healthy colon tissue was then performed using edgeR ^21^. In addition to the differential expression analysis a variety of metrics was calculated for each gene. These metrics included the percentile ranking of each gene’s median expression level in CRC and healthy tissue, and each gene’s area under the curve (AUC) comparing gene expression in CRC tissue to healthy tissue controls. In tables 1 and 2 the differential expression false discovery rate (FDR) and log fold change were calculated using edgeR. The AUC and median percentile ranks in normal/disease tissue was calculated using custom code written in R.

**Table 1:**
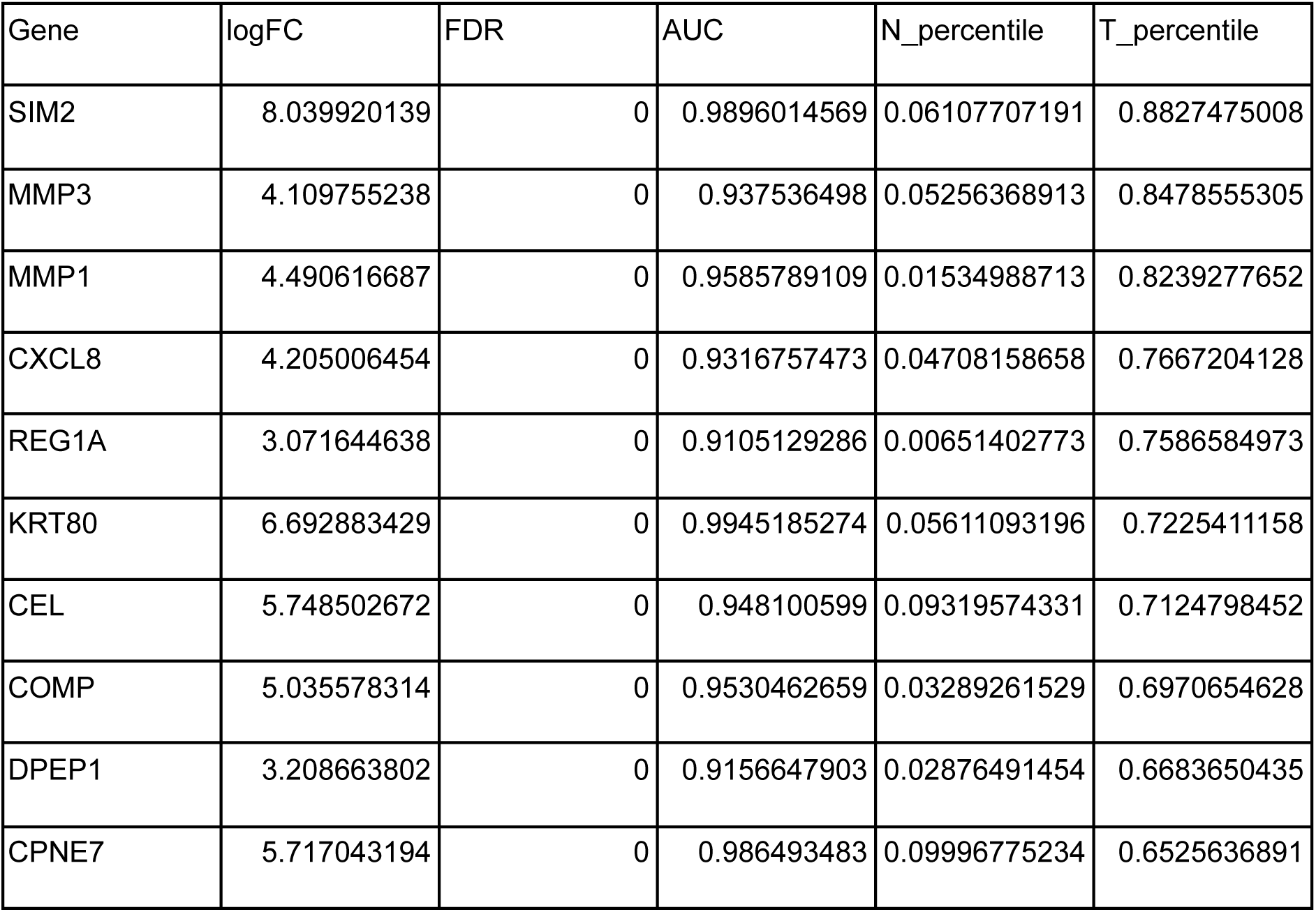
Category 1 Genes. Table 1: *The logFC is the log fold change comparing expression in CRC tissue with normal tissue, FDR is the false discovery rate of the statistical significance of differential expression in CRC tissue compared to healthy colon/rectal tissue. AUC is the area under the curve calculated based on how well the tissue expression can distinguish between CRC and normal colon tissue. The N and T percentile is the percentile ranking of each biomarker’s median expression across all samples in normal or tumor tissue respectively*.

**Table 2:** Category 2 Genes. Table 2: The logFC is the log fold change comparing expression in CRC tissue with normal tissue, FDR is the false discovery rate of the statistical significance of differential expression in CRC tissue compared to healthy colon/rectal tissue. AUC is the area under the curve calculated based on how well the tissue expression can distinguish between CRC and normal colon tissue. The N and T percentile is the percentile ranking of each biomarker’s median expression across all samples in normal or tumor tissue respectively.

| Gene | logFC | FDR | AUC | N_percentile | T_percentile |
| --- | --- | --- | --- | --- | --- |
| TGFB1 | 3.229163005 | 0 | 0.9016781554 | 0.9103514995 | 0.9965172525 |
| RPS10 | 3.647234357 | 0 | 0.9540546642 | 0.5404063205 | 0.9451144792 |
| CEMIP | 5.261626716 | 0 | 0.9846286987 | 0.1985811029 | 0.9361496292 |
| SLCO4A1 | 3.518322484 | 0 | 0.9423060715 | 0.3935504676 | 0.9344727507 |
| MMP11 | 4.389034818 | 0 | 0.9566644592 | 0.3230570784 | 0.932537891 |
| GDF15 | 3.250709446 | 0 | 0.910583667 | 0.3252499194 | 0.9260883586 |
| ETV4 | 5.371444681 | 0 | 0.9864287649 | 0.1584650113 | 0.8612705579 |
| NME2 | 3.644611268 | 0 | 0.9498585232 | 0.4285069332 | 0.8581747823 |
| COL11A1 | 6.461181663 | 0 | 0.9926612685 | 0.1194453402 | 0.8442437923 |
| TRIB3 | 3.834579099 | 0 | 0.9538304085 | 0.2526926798 | 0.8424379233 |

Stool is a complex matrix with microbial RNA being the dominant source of RNA in stool. It is expected that RNA derived from cancer cells will commonly constitute a small fraction of the total RNA present in stool. Therefore additional filters were applied to identify and rank genes in terms of their ability to detect the presence of abnormal cells in stool. We hypothesized that genes with strong differential expression in tissue, comparing CRC tissue samples to normal tissue samples, that are ubiquitously differentially expressed across most/all tumors compared to healthy controls would be good candidates to evaluate for stool based biomarkers applied to CRC/AA screening. In addition, genes that are strongly expressed in CRC tissue are likely easier to detect in stool when the fraction of pathogenic cells is small. The following methodology was used to obtain these genes: differentially expression FDR<0.001 and AUC > 0.9 and a log base 2- fold change difference in expression between CRC and healthy tissue > 2. The genes that passed this filter were then sorted by median expression in CRC samples from highest to lowest. The top of this list would be genes ubiquitously differentially expressed across many CRC tissue samples compared to normal tissue samples that are also highly expressed in CRC tissue. However the genes selected may also be expressed at some level in normal colon tissue as no restriction was placed on expression in tissue from healthy subjects.

In addition to strong differential expression, having low or no expression in normal colon cells would be ideal for a CRC diagnostic biomarker. There are a number of examples of genes with low or no expression in normal colon cells but detectable expression in cancerous tissue ^22,23^. For this category of genes, the ability to detect even a small number of transcripts in stool may be good evidence of the presence of a pathogenic lesion. With this approach, genes were selected using the same set of criteria as outlined above with an additional filter that the percentile ranking of the gene expression in normal colon tissue must be less than 10%. Similar to the first gene set, the genes that passed all filters were sorted by median expression in CRC tissue samples from highest to lowest. The combination of these resultant two gene lists was 158 genes.

The final set of 20 genes tested on clinical samples was selected by taking the top ten genes from each of the gene lists. The top ten genes from the “high expression CRC” gene list are the following: TGFBI, RPS10, CEMIP, SLCO4A1, MMP11, GDF15, ETV4, NME2, COL11A1, TRIB3. The top ten genes from the “low background” gene list are the following: SIM2, MMP3, MMP1, CXCL8, REG1A, KRT80, CEL, COMP, DPEP1, CPNE7. GAPDH was included for use as a reference gene.

### Data Presentation and Statistical Analysis

The delta-delta Ct method ^24^ was used to calculate the relative fold gene expression as plotted in Figure 2. The reference samples used in the delta-delta Ct calculation was the mean Ct value for those cancer samples with detectable signal defined as Ct values < 40 cycles.The reference gene used was GAPDH. For those stool samples with no detectable copies the fold gene expression was set to 0. The machine learning experiments were performed using the tidymodels package in R, default parameters were used for the random forest model. No hyperparameter optimization or feature selection was performed; for the combination of genes, all 20 genes were used as features in the machine learning model, even those genes that individually did not perform well. Occasionally it is observed in the literature performance reported on the same dataset, that hyperparameter optimization and feature selection was performed on. This approach will artificially inflate performance. To get a more accurate measure of classification performance, we only used default parameters and did not tune the machine learning algorithm and did not perform feature selection.

Classification performance was estimated using an average of five-fold cross validations repeated one hundred times with random splits of the data for each cross validation experiment. Five-fold cross validation consists of randomly splitting the data into 5 roughly equal groups, training the model on the combination of four of the groups and testing the model on the held out group. This process is repeated until all groups have been used as the held out test set. The input data to the machine learning experiments was the normalized relative fold-expression for each of the 20 genes. Sensitivity and specificity was calculated using a 0.5 class probability cutoff. If the reported class probability was > 0.5 the sample was predicted to be a disease sample if the class probability was < 0.5 the sample was predicted to be a control sample.

Statistical significance testing differential representation of each of the genes comparing the CRC or AA patient samples with control samples was performed using the Mann-Whitney test, p-values were adjusted for multiple testing comparisons using the Benjamini Hochberg method, FDR <0.05 was used as the significant cutoff. The correlation analysis was performed by calculating the mean counts per million (CPM) for the RNA-Seq datasets across all CRC or normal tissue samples for each of the 20 genes. Then calculating the mean Ct value for each of the 20 genes in the CRC or control stool samples and then calculating the Pearson correlation coefficient comparing CRC tissue with CRC stool samples and comparing normal tissue with control stool samples. Counts per million was calculated using the edgeR package. For those stool samples with undetectable transcripts the Ct value was set to 40 cycles which was the maximum number of cycles used in the qPCR experiments.

### Patients and Sample Collection

From January 2021 to April 2022, a total of 114 clinical fecal samples were collected from patients at Shenzhen Nanshan People’s Hospital. Among them, 33 cases were colorectal cancer (CRC) patients, and 28 cases were colorectal advanced adenoma (AA) patients. The diagnoses of CRC and AA were confirmed through colonoscopic biopsy and pathological examination. Advanced adenomas were defined as adenomas with a diameter >10 mm, or those containing villous components, severe dysplasia, or high-grade intraepithelial neoplasia. The control group consisted of 53 volunteers who underwent health examinations during the same period. Among the controls, 18 cases had negative findings on colonoscopy, while the remaining 35 cases had normal fecal characteristics, negative fecal occult blood tests, and normal results in routine blood tests and tumor marker assessments. All participants were provided with specialized fecal collection tubes before surgery or endoscopic examination. Fresh morning fecal samples (10–15 g) were collected using sampling rods, placed into fecal collection tubes, and sealed. Samples were aliquoted within 2 hours and stored at −80°C for future analysis. Ct values for the biomarkers measured and clinical metadata are listed in supplemental table 1

#### Ethics Approval

This study was approved by the Ethics Committee of Shenzhen Nanshan People’s Hospital (Approval No. KY-2020-045-01). Informed consent was given before stool was collected. The research was performed in accordance with the relevant guidelines and regulations, including the declaration of Helsinki.

### RNA Isolation and PCR amplification

#### RNA extraction

RNA was extracted from 0.25 g of stool using the Omega Bio-Tek E.Z.N.A.® Stool RNA Kit (R6828), which employs a phenol-chloroform extraction method. To remove potential PCR inhibitors, the Zymo Research OneStep™ PCR Inhibitor Removal Kit (D6035) was used following the RNA extraction. The extracted RNA was resuspended in 50 µL of DEPC-treated water per tube. RNA concentration and purity were measured using a NanoDrop spectrophotometer. The typical RNA yield was approximately 450 µg per sample. The RNA was then aliquoted and stored at –80°C until further use.

#### The PCR setup used was as follows

Takara One Step PrimeScript™ III RT-qPCR Mix with UNG (RR601A) was utilized for qRT-PCR. Each reaction was set up with a total volume of 10 µL, including 2 µL of extracted RNA. The reactions were performed on an ABI 7500 Real-Time PCR System with the following thermal cycling conditions: reverse transcription at 50°C for 5 minutes, initial denaturation at 95°C for 10 seconds, followed by 40 cycles of 95°C for 10 seconds and 60°C for 34 seconds.

## Results

We first performed a bioinformatic screen to identify the top 20 genes that based on their expression in tissue (CRC N= 478, Healthy Controls N= 692) were most promising as stool based CRC screening biomarkers. The top 20 genes were grouped into two categories. Category 1 are strongly over expressed genes comparing CRC tissue and healthy colon tissue samples that also have very low expression in healthy colon samples i.e. “low background” genes (See Table 1 and Supplemental Figure 1).

Category 2 are strongly over-expressed genes that had the highest possible expression in CRC tissue i.e. “high signal” genes but which may also have moderate expression in normal colon tissue (See Table 2 and Supplemental Figure 2).

For example RPS10 has very high expression in CRC tissue with a median percentile ranking of 95% in CRC tissue but it is also expressed in normal tissue at an average level with a median percentile ranking in normal tissue of 54%. We then tested the tissue based bioinformatic predictions by measuring the level of representation of these genes in 114 clinical stool samples (CRC N=33, AA N=28, Controls N=53).

We first investigated how well expression in tissue translated to representation in stool. Category 1 genes “low background” genes did not have a higher fraction of genes with no detectable signal in control patients. The fraction of genes comparing Category 1 and Category 2 with undetectable expression in >90% of the stool of control samples was the same at 40% (Figure 2). However, the correlation between the mean Ct value measured in control stool samples and the mean expression level in normal colorectal tissue was consistent with tissue predictions i.e.(high tissue expression had a tendency to have higher expression in stool) and was statistically significant with a Pearson correlation coefficient of –0.57, p-value = 0.007, alpha = 0.05 (Figure 1 panel A). The correlation comparing CRC stool and CRC tissue was similar with a Pearson coefficient of –0.5, p-value = 0.02 (Figure 1 panel B). These results suggest tissue expression has a statistically significant but relatively modest ability to predict the level of gene mRNA representation in stool. We observed with interest that for control samples there was a bias for genes to have lower representation in stool than would be predicted based on tissue expression levels (Figure 1 panel A).

**Fig 1:**
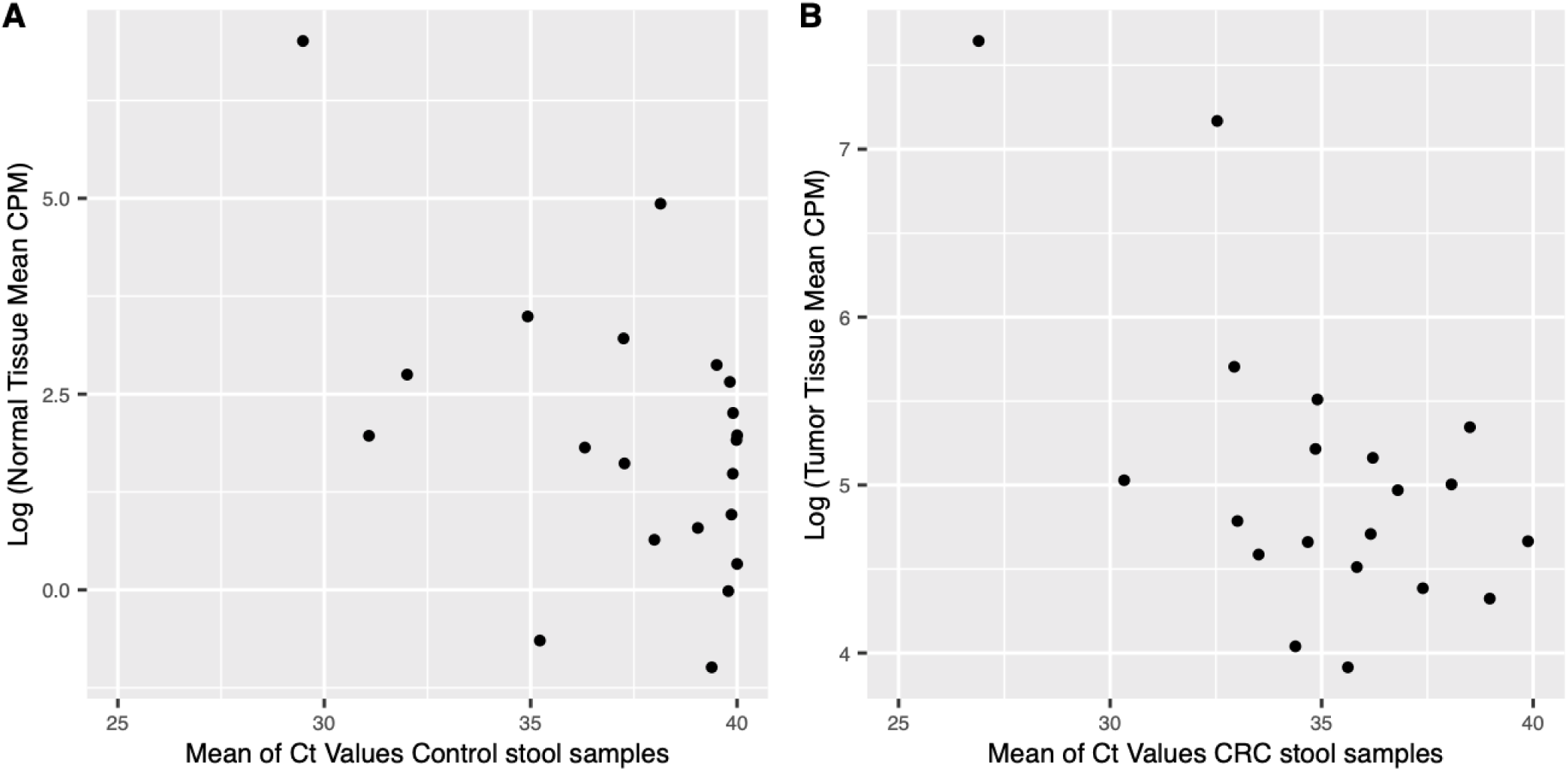
Scatter plot comparing tissue versus stool expression. **Figure 1 Panel A**: Scatter plot Normal tissue versus Control stool (Pearson R= –0.57). Every dot represents a gene. Y-axis is the log of the mean expression across all normal tissue samples, gene expression units are copies per million (CPM), X-axis is the mean Ct value measured across all control stool samples. **Panel B:** Scatter plot CRC tissue versus CRC stool (Pearson R= –0.5). Every dot represents a gene. Y-axis is the log of the mean expression across all CRC tissue samples, gene expression units are copies per million (CPM), X-axis is the mean Ct value measured across all CRC stool samples.

We then looked at how well differential expression observed in tissue was conserved in clinical stool samples. The 20 genes were all strongly overexpressed in CRC tissue compared to normal colon tissue. Of these 20 genes, 14 out of 20 also had statistically significant differential representation comparing CRC stool samples versus control stool samples (Mann-Whitney test, p-values adjusted using the Benjamini Hochberg method, false discovery rate (FDR) <0.05, Table 3). Of the 14 genes that exhibited significant differential expression in CRC relative to normal stool samples, there was a strong trend toward agreement with the tissue analysis. Thirteen (13) of the 14 were over represented in the stool of CRC patients compared to controls, and 1 of the 14 (KRT80) was under represented in the stool of CRC patients compared to controls (Figure 2 and Table 3).

**Figure 2:**
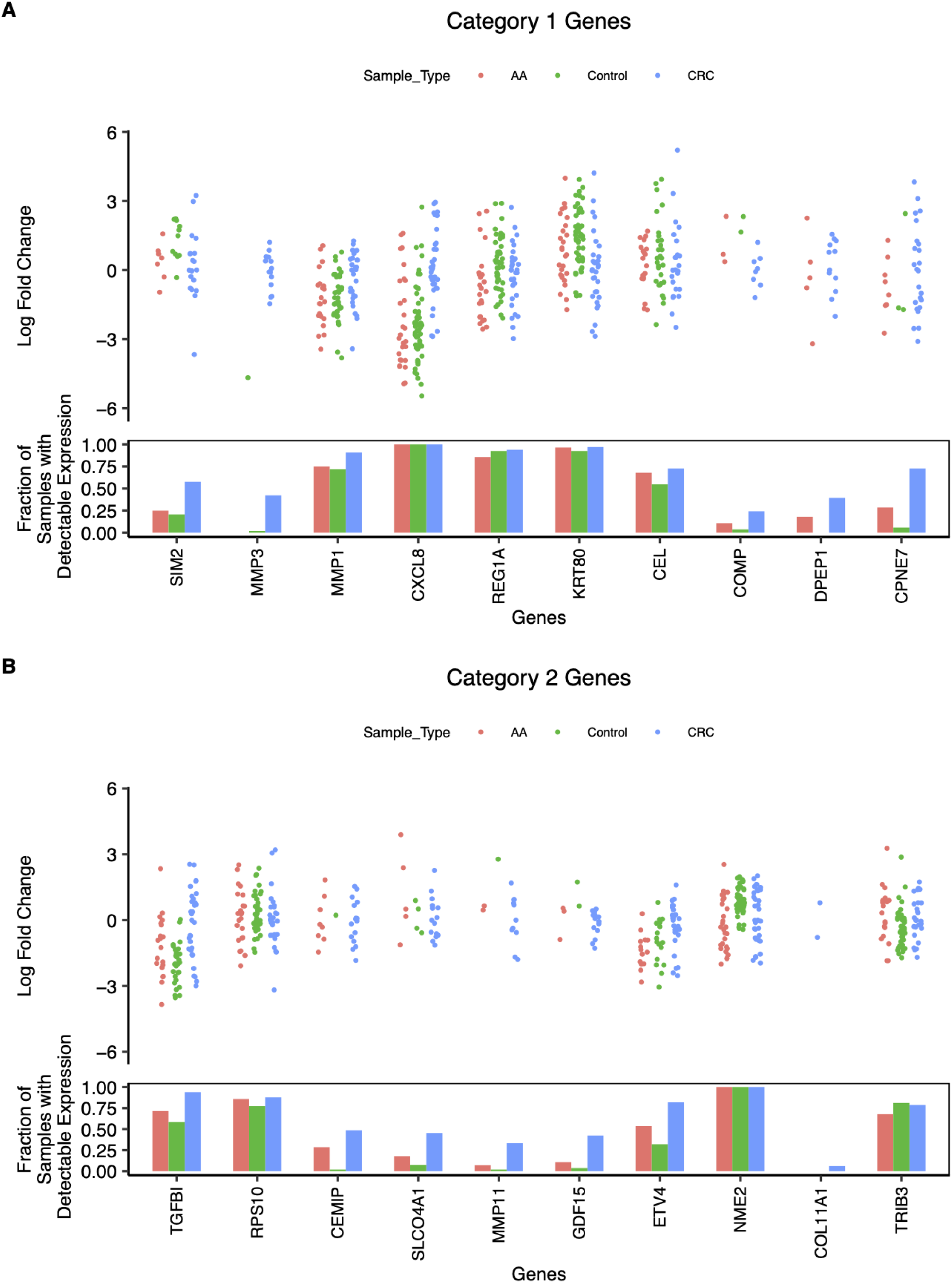
Expression level in clinical stool samples. **Figure 2 panel A**: Plotted normalized representation in stool for category 1 genes i.e. low expression in normal colon/rectal tissue. Every dot represents a sample with a detectable copy number; the top plot is the log fold change calculated using the delta delta Ct method. With the median of CRC samples used as the reference. The bar plot represents the fraction of samples with detectable copy number in stool. **Panel B:** Plotted normalized representation in stool for category 2 genes i.e. high expression in CRC tissue. Every dot represents a sample with detectable transcripts; the top plot is the log fold change calculated using the delta delta Ct method. With the median of CRC samples used as the reference. The bar plot represents the fraction of samples with detectable copy number in stool.

**Table 3:**
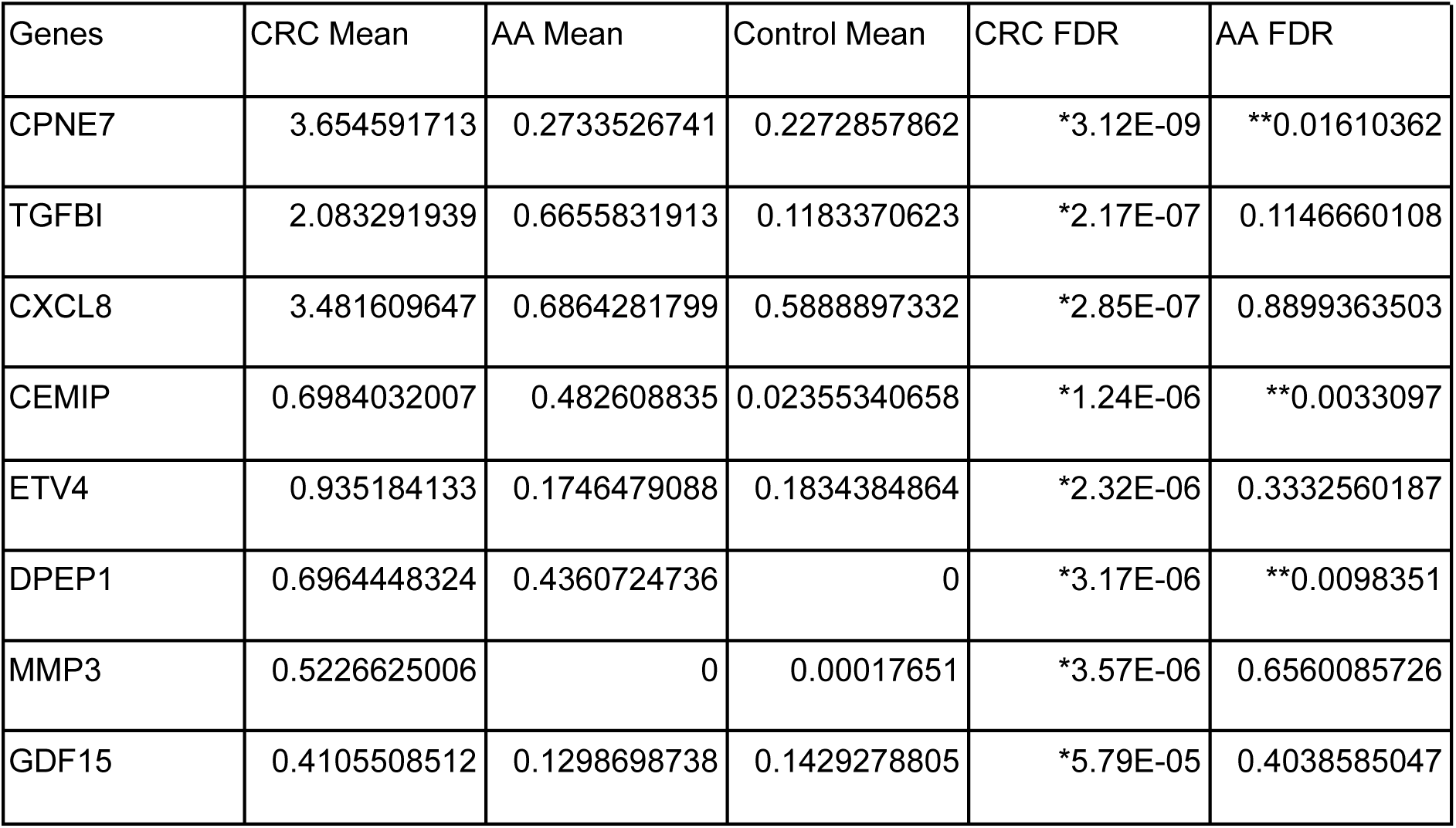
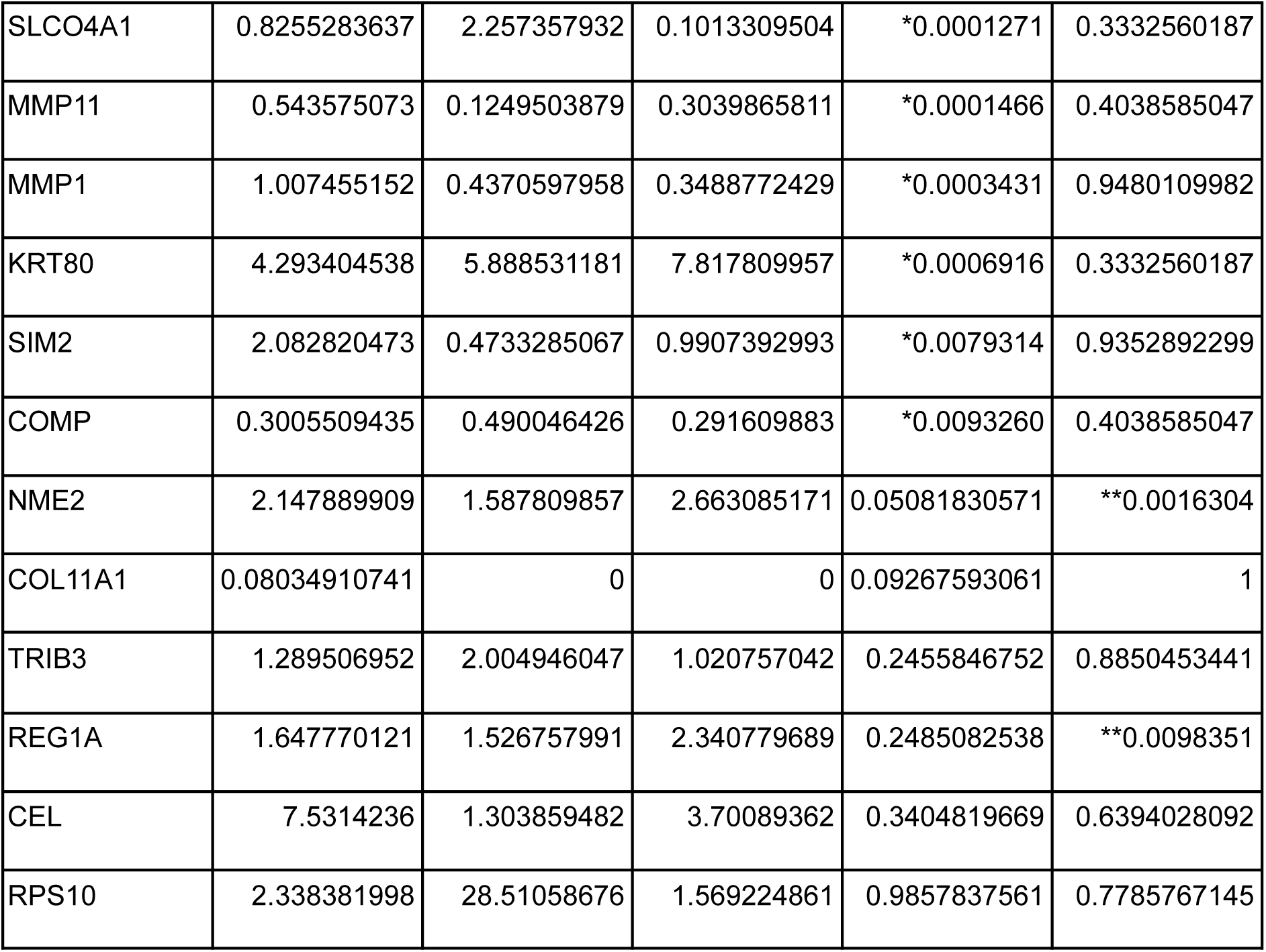
Statistical analysis of gene representation in stool. Table 3: *Mean of relative fold gene expression observed in human stool as calculated using the delta-delta Ct method is given for columns 1 through 3. For samples with no detectable copies their relative fold gene expression was set to zero. Those genes with a single asterisk represent genes with statistically significant differential representation in the stool of CRC patients versus controls. Those genes with a double asterisk represent genes with statistically significant differential representation in the stool of AA patients versus controls. A false discovery rate threshold of 0.05 was used to calculate statistical significance*.

| Genes | CRC Mean | AA Mean | Control Mean | CRC FDR | AA FDR |
| --- | --- | --- | --- | --- | --- |
| CPNE7 | 3.654591713 | 0.2733526741 | 0.2272857862 | *3.12E-09 | **0.01610362 |
| TGFB1 | 2.083291939 | 0.6655831913 | 0.1183370623 | *2.17E-07 | 0.1146660108 |
| CXCL8 | 3.481609647 | 0.6864281799 | 0.5888897332 | *2.85E-07 | 0.8899363503 |
| CEMIP | 0.6984032007 | 0.482608835 | 0.02355340658 | *1.24E-06 | **0.0033097 |
| ETV4 | 0.935184133 | 0.1746479088 | 0.1834384864 | *2.32E-06 | 0.3332560187 |
| DPEP1 | 0.6964448324 | 0.4360724736 | 0 | *3.17E-06 | **0.0098351 |
| MMP3 | 0.5226625006 | 0 | 0.00017651 | *3.57E-06 | 0.6560085726 |
| GDF15 | 0.4105508512 | 0.1298698738 | 0.1429278805 | *5.79E-05 | 0.4038585047 |
| SLCO4A1 | 0.8255283637 | 2.257357932 | 0.1013309504 | *0.0001271 | 0.3332560187 |
| MMP11 | 0.543575073 | 0.1249503879 | 0.3039865811 | *0.0001466 | 0.4038585047 |
| MMP1 | 1.007455152 | 0.4370597958 | 0.3488772429 | *0.0003431 | 0.9480109982 |
| KRT80 | 4.293404538 | 5.888531181 | 7.817809957 | *0.0006916 | 0.3332560187 |
| SIM2 | 2.082820473 | 0.4733285067 | 0.9907392993 | *0.0079314 | 0.9352892299 |
| COMP | 0.3005509435 | 0.490046426 | 0.291609883 | *0.0093260 | 0.4038585047 |
| NME2 | 2.147889909 | 1.587809857 | 2.663085171 | 0.05081830571 | **0.0016304 |
| COL11A1 | 0.08034910741 | 0 | 0 | 0.09267593061 | 1 |
| TRIB3 | 1.289506952 | 2.004946047 | 1.020757042 | 0.2455846752 | 0.8850453441 |
| REG1A | 1.647770121 | 1.526757991 | 2.340779689 | 0.2485082538 | **0.0098351 |
| CEL | 7.5314236 | 1.303859482 | 3.70089362 | 0.3404819669 | 0.6394028092 |
| RPS10 | 2.338381998 | 28.51058676 | 1.569224861 | 0.9857837561 | 0.7785767145 |

Finally, we evaluated the clinical performance of the 20 predicted genes. Both individual and combined area under the curve (AUC) for the 20 genes was calculated using five-fold cross-validation and a random forest classifier ^25^. Comparing CRC versus controls, there were 14 genes with an AUC > 0.6 and 3 with an AUC > 0.8 (CPNE7, TGFBI, CXCL8). The top performing individual biomarker for CRC versus controls was CPNE7 with an AUC of 0.83. The combined performance of all 20 genes, had good synergy with a combined AUC of 0.94 (sensitivity 75%, specificity 95%). In contrast, when compared with the best individually performing gene (CPNE7) with an AUC of 0.83 (sensitivity 69%, specificity 94%, See Table 4, and Figure 3 panel A), clinical performance was enhanced. Hence the combination of genes had considerably better performance than the best performing individual gene.

**Figure 3.**
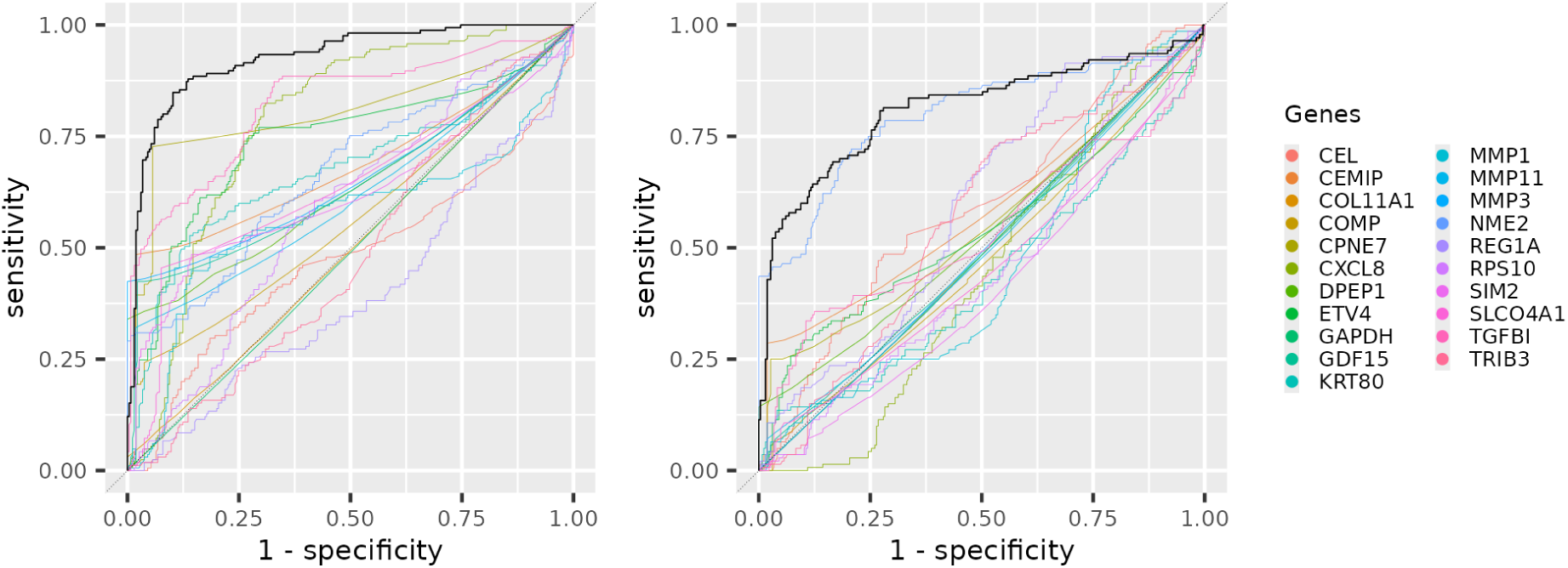
**Panel A:** ROC plot of CRC versus controls, each gene is a different color the combined performance is represented by the solid black line. **Panel B:** ROC plot of AA versus controls, each gene is a different color the combined performance is represented by the solid black line.

**Table 4:** CRC Diagnostic performance of each gene and all genes combined. Table 4: *Area under the curve (AUC) and sensitivity and specificity for each gene comparing CRC versus controls. The performance combining all 20 genes is given in the last row of the table*.

| Genes | AUC | Sensitivity | Specificity |
| --- | --- | --- | --- |
| TGFB1 | 0.8156212121 | 0.62 | 0.8150909091 |
| RPS10 | 0.577491342 | 0.3452380952 | 0.7352727273 |
| CEMIP | 0.7283463203 | 0.4819047619 | 0.9810909091 |
| SLCO4A1 | 0.6803571429 | 0.4385714286 | 0.9247272727 |
| MMP11 | 0.6496623377 | 0.3219047619 | 0.9810909091 |
| GDF15 | 0.6944090909 | 0.4257142857 | 0.9756363636 |
| ETV4 | 0.753508658 | 0.5752380952 | 0.8267272727 |
| NME2 | 0.675478355 | 0.4580952381 | 0.7550909091 |
| COL11A1 | 0.5102380952 | 0.02047619048 | 1 |
| TRIB3 | 0.5247835498 | 0.319047619 | 0.7170909091 |
| SIM2 | 0.6837012987 | 0.3804761905 | 0.9054545455 |
| MMP3 | 0.7121428571 | 0.4242857143 | 1 |
| MMP1 | 0.5456277056 | 0.4842857143 | 0.7816363636 |
| CXCL8 | 0.8313766234 | 0.7123809524 | 0.7661818182 |
| REG1A | 0.413512987 | 0.2380952381 | 0.6463636364 |
| KRT80 | 0.6934004329 | 0.5257142857 | 0.8076363636 |
| CEL | 0.4716017316 | 0.2971428571 | 0.7812727273 |
| COMP | 0.6023441558 | 0.240952381 | 0.9623636364 |
| DPEP1 | 0.68 | 0.36 | 1 |
| CPNE7 | 0.8346233766 | 0.6876190476 | 0.9436363636 |
| Combined Genes | 0.9431558442 | 0.7547619048 | 0.9510909091 |

Six of the genes with an AUC > 0.6 exhibited very good specificity (>0.95%) with reasonable sensitivity (24%-48% sensitivity) (Table 4, CEMIP, MMP11, GDF15, MMP3, COMP, DPEP1) and hence would be excellent candidates to combine together in a panel. Two of the genes (MMP3, DPEP1) had close to 100% specificity, DPEP1 was not detected in any of the 35 control samples but was detected in 36% of the CRC samples and 18% of the AA samples. MMP3 was detected with a low copy number in 1 of the control samples (3% of controls) but had detectable copies in 42% of the CRC samples). Highly specific genes like these would be excellent candidates to combine as part of a clinical diagnostic panel.

In contrast with the CRC samples relative to control subjects the majority of genes were not significantly differentially represented in the stool of AA patients compared to the control subjects. Of the 20 genes 5 out of 20 had statistically significant differential representation comparing AA stool samples versus control stool samples (Mann-Whitney test, p-values adjusted using the Benjamini Hochberg method, false discovery rate (FDR) <0.05, Table 3, CPNE7, CEMIP, DPEP1, NME2, REG1A), with 4 of the 5 being over represented in the stool of AA patients. Of the five genes NME2 was the only gene that was under-represented in the stool of AA patients. The direction of representation comparing AA with controls was consistent with the CRC results. The 4 genes that were over represented in the stool of AA patients were also over represented in the stool of CRC patients. And the one gene (NME2) that was underrepresented in the stool of AA patients was also under-represented in the stool of CRC patients. The best performing gene discriminating between AA and control stool samples was NME2 with an AUC of 0.82. In contrast to the CRC results the combined performance of all 20 genes did not show much synergy with an AUC of 0.83 which was approximately the same as the best performing AA gene (sensitivity 56%, specificity 93%, See Table 5, and Figure 3 panel B).

**Table 5:** AA Diagnostic performance of each gene and all genes combined. Table 5: *Diagnostic performance for each gene comparing AA versus controls. The performance combining all 20 genes is given in the last row of the table*.

| Genes | AUC | Sensitivity | Specificity |
| --- | --- | --- | --- |
| TGFB1 | 0.5833939394 | 0.39 | 0.8474545455 |
| RPS10 | 0.4970727273 | 0.262 | 0.732 |
| CEMIP | 0.6287272727 | 0.276 | 0.9814545455 |
| SLCO4A1 | 0.5057939394 | 0.09533333333 | 0.962 |
| MMP11 | 0.5258787879 | 0.07066666667 | 0.9810909091 |
| GDF15 | 0.5099575758 | 0.03933333333 | 0.9625454545 |
| ETV4 | 0.5549272727 | 0.3113333333 | 0.8447272727 |
| NME2 | 0.8178787879 | 0.54 | 0.8983636364 |
| COL11A1 | 0.5 | 0 | 1 |
| TRIB3 | 0.6398545455 | 0.408 | 0.7287272727 |
| SIM2 | 0.4682606061 | 0.05466666667 | 0.9263636364 |
| MMP3 | 0.5 | 0 | 1 |
| MMP1 | 0.4731151515 | 0.1906666667 | 0.7925454545 |
| CXCL8 | 0.4855818182 | 0.224 | 0.728 |
| REG1A | 0.6231212121 | 0.3686666667 | 0.6956363636 |
| KRT80 | 0.4268848485 | 0.1866666667 | 0.7309090909 |
| CEL | 0.6239515152 | 0.316 | 0.8094545455 |
| COMP | 0.4994363636 | 0.054 | 0.9638181818 |
| DPEP1 | 0.5713333333 | 0.1426666667 | 1 |
| CPNE7 | 0.6020121212 | 0.2546666667 | 0.9683636364 |
| Combined Genes | 0.8265212121 | 0.558 | 0.9265454545 |

## Discussion

A systematic and comprehensive investigation of mRNA biomarkers for the purposes of CRC screening has not yet been done. In this proof of concept study we have partially filled this gap. We performed a bioinformatic screen of public tissue transcriptomics datasets and ranked genes according to suitability for stool based screening. We then evaluated how well this tissue based analysis is predictive of transcript representation in stool.

Our findings indicate that tissue expression profiles are modest predictors of stool transcript abundance but are good predictors of which genes are likely to be differentially represented in the stool of CRC patients. The correlation between tissue and stool transcript levels was only moderate with a correlation coefficient of ∼ –0.5. However in agreement with the tissue results, the majority of genes were differentially represented in the stool of CRC patients compared to controls. And the direction of representation had strong concordance with the tissue based prediction. Of the 14 out of 20 genes that were significantly differentially represented in stool, 13 of them were overrepresented in CRC patients, which matched the direction of expression predicted by the tissue analysis. The exception was KRT80 with statistically significant lower representation in the stool of CRC patients compared to controls (Table 3). It is unclear why KRT80 is the lone exception. But it may have something to do with the suspected role KRT80 plays in regulating apoptosis ^26^. Normal cells detached from the extracellular matrix and shed into the lumen commonly undergo apoptosis ^27^) so genes involved in regulating apoptosis may well be differentially expressed in normal cells shed into the lumen compared to expression in tissue.

Overall tissue predictions performed relatively well at predicting which genes will be differential represented and the direction of the differential representation i.e. over or under represented in CRC compared to control stool samples. Tissue results were mediocre at predicting the *degree* of differential representation and performed poorly at predicting which genes would have low or no representation in control stool samples. In particular there was a bias for transcript levels measured in control stool to be lower than would be expected based on their expression in normal colon tissue (See Figure 1 panel A). This bias for genes to have a lower representation in stool may be a combination of stress response pathways being activated in shed cells and shed normal colon cells being enriched for apoptotic cells^27^; stress response and apoptosis leads to general repression of gene transcription^28^. Cancer cells in contrast are resistant to apoptosis^29^. It has been shown that long DNA fragments presumably representing non apoptotic cells are enriched in the stool of CRC patients ^30^.

The tissue based predictions performed well at predicting genes likely to have clinical utility. More than half the genes identified in the in silico tissue screen were differentially represented in the stool of CRC patients with 3 of them having an AUC > 0.8 (TGFBI, CXCL8, CPNE7) and a combined AUC using all 20 genes of 0.94 (sensitivity 75%, specificity 95%). Unsurprisingly, the AA prediction performance was somewhat lower than the CRC performance with an AUC of 0.83 (sensitivity 55.8%, specificity 92.6%) but the AA performance compares very well with published AA performance for nucleic acid based biomarkers^16,31^. And is a good deal better than published FIT AA detection rates^11^.

Many of the genes had very low transcript numbers in control samples. One gene (DEP1) had no detectable expression in any of the control samples and for three genes (MMP3, MMP11 and CEMIP) only one of the control samples had detectable expression. This suggests a straightforward way to improve diagnostic performance would be to improve assay sensitivity perhaps by increasing the amount of stool processed and subsequent RNA extracted. Ideally this would increase the sensitivity of detection of pathogenic lesions without compromising the specificity.

Of the 20 genes tested only one of them to our knowledge (DPEP1) has been tested before as a mRNA stool based biomarker for CRC screening ^32,33^. Therefore most of the genes that have been tested in the literature do not represent the most promising candidates based on tissue expression. In contrast, we tested the most promising genes based on tissue analysis. Our study adds considerably to the pool of promising mRNA biomarkers published in the literature.

Limitations of this study include biased sample collection (enriched for rectal cancers, see supplemental table 1), small clinical sample size, older samples in some cases ∼2 years old and a relatively small set of genes tested on clinical samples. In particular 18 of the samples in the control group were colonoscopy negative; the rest of the control group were volunteers without colonoscopy negative confirmation. Given the low incidence rate of colorectal cancer in the general population it is unlikely the control group included a patient with undiagnosed colorectal cancer. But given the prevalence of AAs it is likely undiagnosed AAs were present in the control group. Undiagnosed pre-cancerous lesions may have caused some genes to appear less specific than they truly are since any samples with a high level of signal in the control group were treated as a false positive in the analysis.

While the cohort of clinical samples was relatively small, all the genes tested in the clinical cohort were first screened for strong differential expression, using a large high quality cohort of tissue samples. In general the clinical results were in agreement with the larger tissue based analysis. The bioinformatic screen using tissue samples predicted more than 150 genes with a high likelihood of being useful CRC biomarkers. In this preliminary study we tested only the top 20 genes, given the promising results further testing of additional genes is in progress.

## Conclusions

RNA based biomarkers are a promising approach to CRC and AA stool based screening. However, individually testing genes in clinical stool samples to identify which mRNA biomarkers to prioritize is a laborious and expensive process. Here we have demonstrated that a bioinformatic screen based on tissue expression profiles can productively identify genes with promising clinical utility for stool based CRC and advanced precancerous lesion diagnostics. We tested the top 20 bioinformatically ranked genes and the majority of them had statistically significant differential representation in the stool of CRC patients compared to controls. And many of them are novel mRNA biomarkers that have strong diagnostic performance. In addition we show that expression in tissue does have some predictive power for expression in stool.

The ability to quickly identify promising stool based biomarkers on the basis of the large number of high quality publicly available tissue transcriptomics datasets greatly facilitates the identification of mRNA biomarkers with strong clinical utility.

## Additional Information

### Authors’ contributions

*Concept and Clinical Study Design-* Dr. Du, Wenying Pan, Loren Hansen, Houcong Liu, Xiao Yang

*Patient recruitment and sample collection:* Houcong Liu, Zhufang Chen, Hekai Zhou, Dr Du

*Data Analysis*: Loren Hansen, Wenying Pan

*Drafting of the Manuscript*: Loren Hansen, Houcong Liu

*Wet lab method development and processing samples*: Changpu Song, Haijiu Lin, Dan Chen

*Critical review of the manuscript for important intellectual content:* All authors.

### Availability of data and materials

Clinical data including measured Ct values for each gene and sample is contained in supplemental table 1.

### Consent for Publication

All authors have approved the manuscript for submission. And the content of this manuscript has not been published elsewhere.

## Data Availability

All data produced in the present study may be available upon request to the the authors.

